# A five-day Mindfulness Based Intervention to reduce burnout and psychological distress in UK Medical Students

**DOI:** 10.64898/2026.11.03.26367152

**Authors:** Anne Crombie, Manmeet Kaur, Ned Powel, Menna Brown

## Abstract

**Background:** Medical professionals face significant work overload, with excessive job demands often strongly associated with attrition and burnout. Consequences of burnout include reduced job satisfaction, absenteeism, reduced patient care, lower patient satisfaction and increased healthcare costs. UK Medical students are also at high risk of burnout. Mindfulness-based interventions are effective individual-level treatments; however, more research is required to investigate the efficacy for healthcare professionals and students.

**Method:** Four cohorts of second-year medical students at Cardiff University participated in a five-day, in-person mindfulness-based intervention. Pre (T1) and post (T2) intervention and follow-up (T3) survey data were collected online.

**Results:** All outcome measures saw a statistically significant improvement (N=69) post intervention, current energy, calmness, focus and stress (all P=0.001); Psychological distress (Core-10, P=0.001), Burnout (OLBI, P=0.023), and Wellbeing (WHO5, P=0.37) with small, large and small effect sizes respectively; maintained at follow-up. Satisfaction ratings were consistently high, and completion rate was 100%.

**Discussion:** Medical schools should consider implementing mindfulness-based interventions into medical curricula, as a feasible and acceptable student selected preventative tool to address burnout and incorporate student perspectives on delivery; to prevent mental ill health in medical professionals.

## Background/ Introduction

The global burden of mental ill health is significant, accounting for approximately 13% of the total global burden of disease. Mental illness is now the leading cause of disability worldwide.^1^ UK estimates suggest approximately 20% of the population have a common mental health disorder (CMD) with an associated financial burden of £117.9 billion in 2019.^2^ Rates are significantly higher for medical professionals.^3^ This includes the incidence of burnout, reported rates of which exceed 50%.^4^

Burnout arises from chronic workplace stress that has not been successfully managed^5^, often classified by, energy depletion or exhaustion, cynicism and decreased professional efficacy.^6^ Burnout causes severe depersonalisation and lack of belief in one’s professional capability ^7^, which can detrimentally impact self-esteem and ability to function. The syndrome often develops gradually, meaning that early signs could be disregarded as stress or a busy work period, potentially minimising the opportunity for preventative intervention.^8^

Burnout occurs mainly in the caring professions, medics, nurses, social workers, and teachers. ^9^ A recent NHS staff survey reported that over 30% of clinical staff had experienced burnout due to systemic pressures^10^. Consequences for medical professionals are well documented and include reduced job satisfaction, absenteeism, cynicism, poor self-care, reduced patient care, lower patient satisfaction and increased healthcare costs. ^9^

Initial predictors of burnout may originate during medical school.^11^ Medical students experience high prevalence of CMD compared to age-matched peers studying alternative courses.^12^ Outcomes often worsen throughout medical education, with some students showing poorer emotional health profiles by the end of their degrees.^13^ Several factors are thought to contribute to poor medical student wellbeing, such as excessive workload and compromised work life balance, financial stressors and assessment-related performance pressures.^14^ Burnout in medical students is comparable to medical professionals^11,15^ and high levels of burnout are associated with decreased life satisfaction and serious thoughts of dropping out; leading to poor academic performance, reduced commitment and higher levels of stress. ^16^ In some cases, persistent burnout is associated with psychiatric disorders and suicidal ideation.^15^ It is therefore vital for medical students to have access to effective mental health and wellbeing support.

Several barriers prevent medical students from receiving or engaging with such support, including a fear of negative effect on career opportunities, concern of confidentiality breach, shame from peers and lack of time alongside academia.^17^ Recently, UK medical schools have begun to actively support students’ wellbeing, via wellness interventions and education programmes which incorporate self-care skills.^18^ Ideally, interventions are integrated into pre-existing curricula, to promote positive wellbeing and act as a preventative system for CMD and burnout.^11^

Mindfulness-based interventions (MBIs) offer an evidence-based support, effective for depression^19^, anxiety^20^, pain management and quality of life^21^, persistent at six-month follow-up^22^. Over recent years they have received increased attention within a medical context with positive outcomes for healthcare professionals reported for example ^23, 24^. Mindfulness is a non-judgemental awareness and observation of internal and external events as they arise, in the present moment.^25^ All MBIs include mindfulness and mindfulness-based activities, but there is variation within structured programmes in content, duration and delivery mode. For example, some MBIs apply Sudarshan Kriya Yoga (SKY) breathing; an evidence-based approach^26^ associated with significant improvements across six mental health domains^27^, improved lipid profiles and haematological parameters in students undergoing exam stress^28^ with potential to combat burnout in medical students.^18,29^

### Aim

Thus, this service evaluation aimed to assess the feasibility and acceptability of an MBI within a Welsh (UK) undergraduate medical curricular, as an optional module; to evaluate outcomes for burnout, psychological distress and subjective wellbeing.

## Method

### Study Design

Service evaluation of an MBI using pre, post and follow assessment.

### Ethics

Cardiff University School of Medicine Research Ethics Committee (REC) reviewed the study documents and proposal and considered the project to be service evaluation and under their guidelines confirmed that ethical approval was not required.

### Intervention

The MBI was led by Dr Manmeet Kaur and supported by trained facilitators from the Unity in Health Cymru CIC team, who assisted with practical sessions, group activities, participant support and programme logistics.

The SKY Breathing programme is a structured breathing intervention that combines guided rhythmic breathing exercises with meditation and relaxation techniques to promote mental wellbeing, resilience and stress reduction.^30^

The Holistic Health Care (HHC) SKY breathing Programme was delivered in person over five consecutive days at the University Hospital of Wales. Each session lasted approximately three hours and combined interactive teaching, experiential learning, practical skills and reflective practice. An overview of the programme content is presented in Table 1.

**Table 1:**
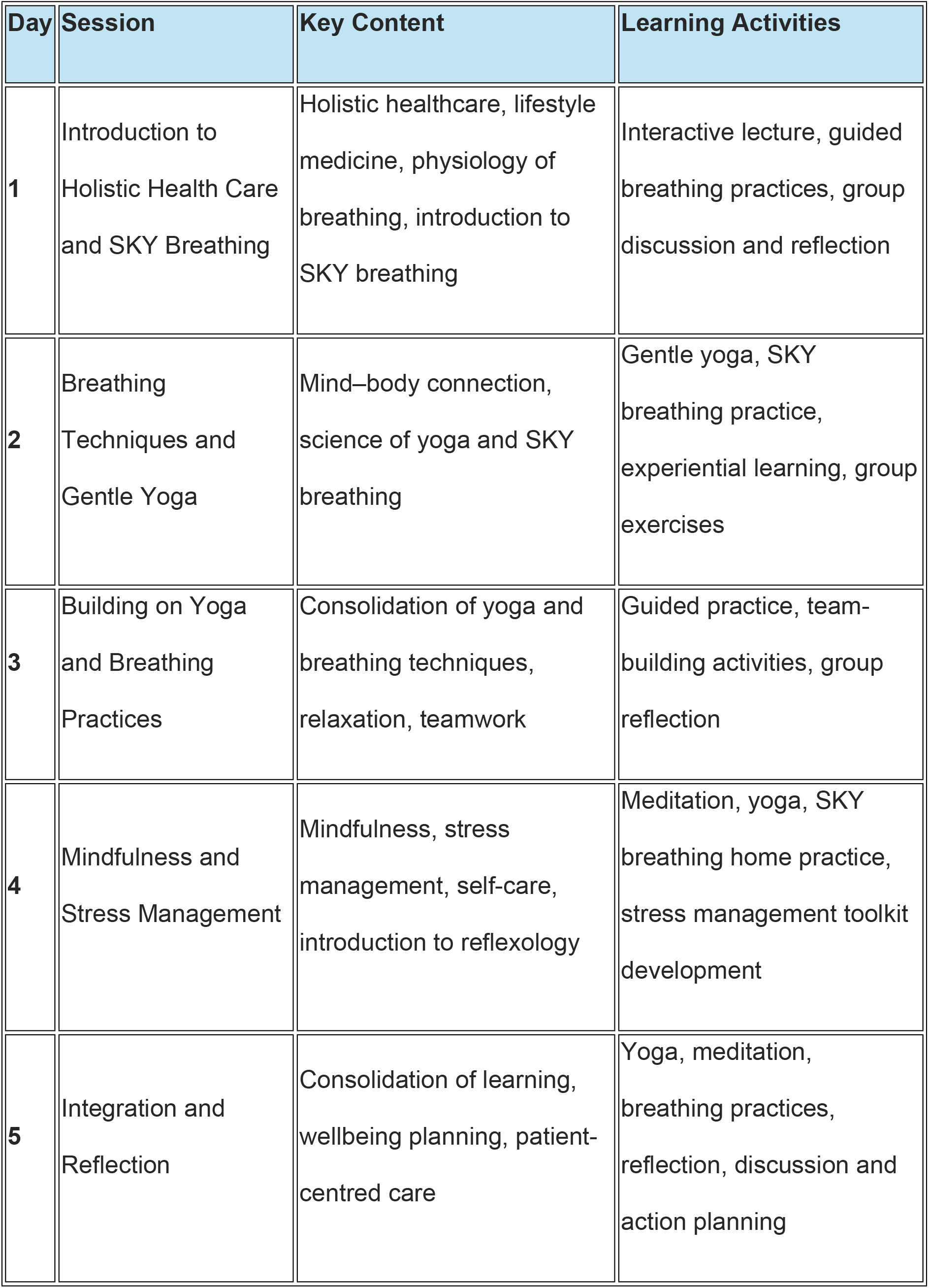
HHC SKY breathing programme - content overview.

| **Day** | **Session** | **Key Content** | **Learning Activities** |
|---|---|---|---|
| **1** | Introduction to Holistic Health Care and SKY Breathing | Holistic healthcare, lifestyle medicine, physiology of breathing, introduction to SKY breathing | Interactive lecture, guided breathing practices, group discussion and reflection |
| **2** | Breathing Techniques and Gentle Yoga | Mind–body connection, science of yoga and SKY breathing | Gentle yoga, SKY breathing practice, experiential learning, group exercises |
| **3** | Building on Yoga and Breathing Practices | Consolidation of yoga and breathing techniques, relaxation, teamwork | Guided practice, team-building activities, group reflection |
| **4** | Mindfulness and Stress Management | Mindfulness, stress management, self-care, introduction to reflexology | Meditation, yoga, SKY breathing home practice, stress management toolkit development |
| **5** | Integration and Reflection | Consolidation of learning, wellbeing planning, patient-centred care | Yoga, meditation, breathing practices, reflection, discussion and action planning |

### Participants

Participants were second year medical students, aged 18 plus, currently enrolled on a five-year, taught, undergraduate medical programme at Cardiff University School of Medicine.

### Procedure

The MBI was offered as a Student Selected Component (SSC) within the medical curriculum. SSCs were advertised and students self-selected their preferred programme.

Following self-selection of the MBI students were invited via email to attend a pre-intervention online briefing delivered by Dr Kaur. The invitation email included the Participant Information Sheet, consent form and baseline (T1) survey. Immediately following the final session participants were invited by email to complete the post-intervention (T2) survey. This survey replicated the baseline measures and included additional questions evaluating participant satisfaction. A follow-up survey (T3) was emailed three months after programme completion to assess the sustainability of outcomes.

Programme fidelity was monitored throughout the intervention using daily attendance registers and programme delivery records maintained by Dr Kaur. Anonymised data were collated and analysed independently from the delivery team.

#### Outcome measures

All surveys included three validated outcome measures;

### Oldenburg Burnout Inventory (OLBI)

The OLBI is a validated 16-item tool measuring burnout through two sub-scales; exhaustion (feelings of tiredness or weariness) and disengagement (detachment and negative attitudes towards work). Responses are categorised using a 4-point scale (Strongly Agree to Strongly Disagree). Scores are presented as a total score (range of 16-64); higher scores indicate a higher level of burnout^31^ and have been classified as low, moderate or severe burnout.^32^

### Clinical Outcomes in Routine Evaluation 10 (CORE-10)

The CORE-10 is a validated ten-item measure that explores individuals’ feelings over the past week, measuring anxiety, depression, trauma, physical problems, functioning and risk to self. Higher scores (range 0-40) indicate greater distress and are classified into six severity levels ranging from heathy to severe. Scores above 11 indicate clinically significant distress.^33^

### World Health Organisation 5 Wellbeing Index (WHO-5)

The WHO-5 measures subjective wellbeing.^34,35^ It consists of five positively phrased statements evaluating mood, vitality, and general interest over the preceding two-week period, rated on a 6-point scale. Scores are summed, (range 0-25) and converted to percentages. Higher scores indicate positive wellbeing, a score below 50 (or a raw score below 13) is considered a cut-off for poor mental wellbeing and suggests further assessment is required.^35^ This measure was added for cohorts two onwards.

### Demographic, feasibility and satisfaction items

Age and gender were recorded at baseline (T1); participants indicated reasons for selecting the MBI from a list of five options. Openness to using non-drug interventions in daily life and in medical practice was measured using a 5-point Likert scale. As were current energy, stress, sustained focus, and calmness.

Post intervention (T2) participants were asked about their satisfaction with the programme, the instructors (using a four-point Likert scale), and whether they would recommend the programme to others (yes/no), with opportunity for free text comments.

### Data analysis

Numerical data was analysed in SPSS. Descriptive analysis calculated Mean (M) scores, standard deviation (SD), number (N) and frequencies. Paired samples t-tests identified statistically significant differences, after ensuring data were normally distributed with equal variance. Follow-up data were analysed using Wilcoxen signed ranks. Effect sizes were calculated and Cohen’s d interpretation applied.^36^ Completion rate was calculated as a percentage. Intervention delivery was assessed by discussing (AC), with Dr Kaur, changes made to delivery at the end of each programme, reported narratively.

## Results

### Participant summary

Data from 69 participants across four cohorts were analysed. The majority were female (43/62, 62.3%), mean age was 20.10 years (SD=1.051), range 19-23 (Table 1).

**Table 1:** Demographic characteristics of all HHC intervention survey respondents.

| **Demographic** | N | % |
|---|---|---|
| **Cohort** |  |  |
| One | 16 | 23.1 |
| Two | 16 | 23.1 |
| Three | 17 | 24.6 |
| Four | 17 | 24.6 |
| **Gender** |  |  |
| Female | 43 | 62.3% |
| Male | 19 | 27.5% |
| Non-binary | 0 | / |
| Prefer not to say | 0 | / |
| Did not report | 7 | 10.1% |
| **Age** |  |  |
| 19 | 20 | 28.9% |
| 20 | 25 | 36.2% |
| 21 | 10 | 14.4% |
| 22 | 5 | 7.2% |
| 23 | 2 | 2.8% |
| Missing | 7 | 10.1% |
| **Total** | **69** |  |

### Bespoke survey items

Participants in all cohorts self-reported current energy and stress. Participants in cohorts two onwards also self-reported sustained focus and calmness. All four measures saw statistically significant improvement post intervention (T2) (P=.001) (Table 2).

**Table 2:** Table of comparison of energy, stress, ability to sustain focus and calmness levels pre (T1) and post (T2) intervention. Mean values, paired t-test, and p-values are presented for each measurement (SD = standard deviation).

| **Measurement** | **N** | **Mean<br>Pre** | **Mean<br>Post** | **SD<br>Pre** | **SD<br>Post** | **p-value** |
|---|---|---|---|---|---|---|
| Energy | 63 | 3.05 | 3.79 | 0.906 | 0.859 | 0.001 |
| Stress | 63 | 3.35 | 2.20 | 0.919 | 0.997 | 0.001 |
| Sustain focus | 47 | 2.70 | 3.45 | 0.749 | 1.017 | 0.001 |
| Calmness | 47 | 2.72 | 4.22 | 0.971 | 0.814 | 0.001 |

Participants demonstrated considerable openness to using non-drug interventions in daily life and in medical practice; 36.2% (25/46) selected ‘very *open*’, 24.6% (17/46) *‘open*’, and 7% (4/46) ‘*neutral*’. The majority of participants reported the following reasons for self-selecting the MBI: ‘*to improve my mental health’* (28/49, 57.1%) and ‘*I was curious’* (27/49, 55%).

### Outcome measures

Participants in all cohorts completed the OLBI and CORE-10. Participants in cohorts two onwards also completed the WHO-5. Participants saw a statistically significant improvement post intervention (T2) for burnout (OLBI, P=.02), psychological distress (CORE-10, P=.00) and wellbeing (WHO-5, p=.03) (Table 3) with, small (.364), large (.983) and small (.280) effect sizes respectively (using Cohens d).

**Table 3:** Comparison of mean scores, standard deviation and p-values for CORE-10, WHO-5 and OLBI pre (T1) and post (T2) intervention, calculated using Paired samples t-test.

| **Measurement** | **N** | **Mean T1** | **Mean T2** | **SD T1** | **SD T2** | **p-value** |
|---|---|---|---|---|---|---|
| CORE-10 (Psychological Distress) | 63 | 11.16 | 6.50 | 5.793 | 4.474 | 0.001 |
| OLBI (Burnout) | 62 | 39.06 | 36.81 | 5.031 | 5.655 | 0.005 |
| WHO-5 (raw score) (Wellbeing) | 47 | 15.60 | 17.33 | 2.909 | 4.153 | 0.037 |

### Oldenburg Burnout Inventory (OLBI)

At both T1 and T2 the majority of the cohort reported ‘low levels’ of Burnout (54/62, 87% and 57/59, 96.6% respectively). Post intervention burnout severity reduced at all levels (Table 4).

**Table 4:**
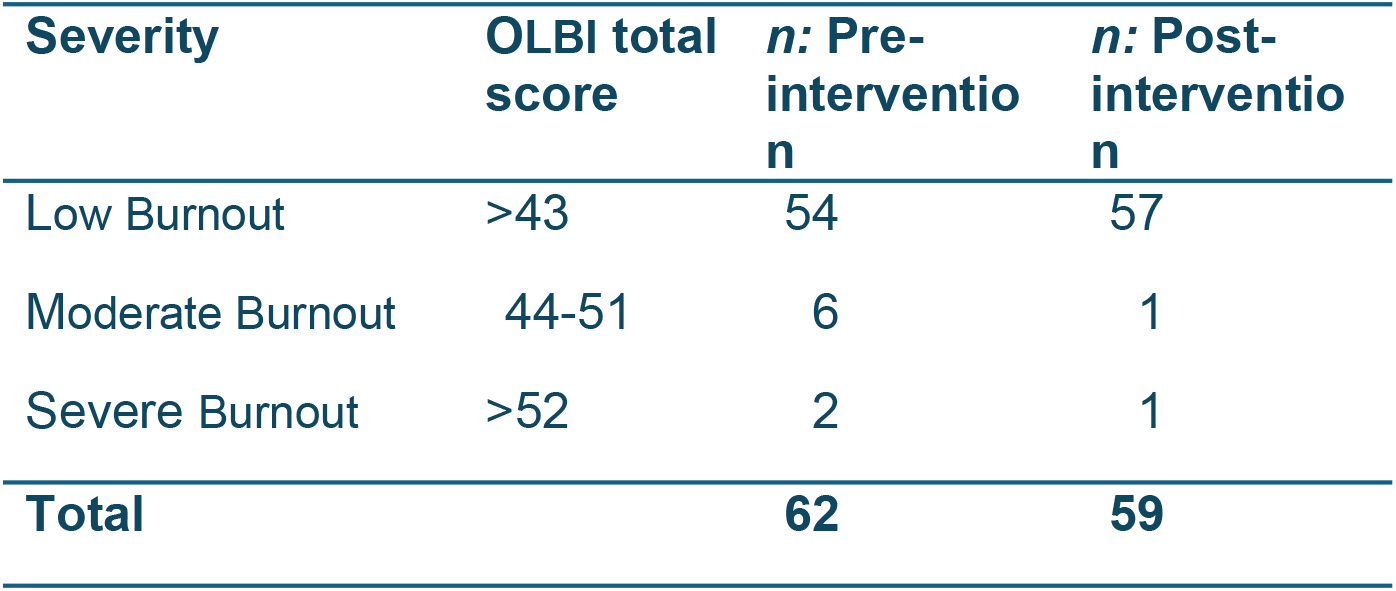
Table of OLBI total scores pre (T1) and post (T2) intervention with corresponding severity of Burnout and percentages reported.

The OLBI includes two sub scales: Exhaustion, where higher scores (23 +) indicate greater physical, affective and cognitive fatigue; and, disengagement, where higher scores (22 +) reflect a stronger tendency for individuals to distance themselves from their work and develop negative attitudes toward their job. Table 5 shows a statistically significant decrease in the total mean exhaustion.

**Table 5:**
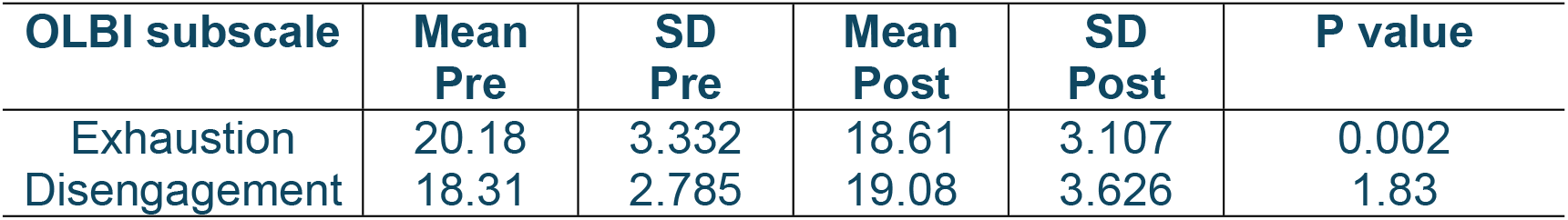
Table of Exhaustion and Disengagement sub-scores pre intervention (T1) and post intervention (T2). Mean values, paired samples t-test, and p-values are presented for each measurement (SD = standard deviation), N=55.

| **OLBI subscale** | **Mean Pre** | **SD Pre** | **Mean Post** | **SD Post** | **P value** |
|---|---|---|---|---|---|
| Exhaustion | 20.18 | 3.332 | 18.61 | 3.107 | 0.002 |
| Disengagement | 18.31 | 2.785 | 19.08 | 3.626 | 1.83 |

### CORE-10 - Psychological distress

At baseline (T1) the combined cohort data indicated a range of psychological distress levels, with the majority meeting the threshold for ‘low level distress’ (19/63, 30%) and/or ‘mild’ distress (18/63, 28.5%). A small percentage self-reported within the healthy range (12/63, 19%). At T2 the majority of participants reduced their distress and entered the ‘healthy’ range (39/60, 60%). Table 6 shows the decrease in severity of psychological distress post intervention.

**Table 6:** Table presents the number of participants meeting each Core-10 severity category pre (t1) and post-intervention (T2) with percentages reported.

| **Severity** | **CORE-10 total score** | ***n:* Pre-intervention** | ***%*** | ***n:* Post-intervention** | ***%*** |
|---|---|---|---|---|---|
| Healthy | 0-5 | 12 | 19.0 | 39 | 65.0 |
| Low level | >5-10 | 19 | 30.1 | 19 | 31.6 |
| Mild | >10-15 | 18 | 28.5 | 9 | 15.0 |
| Moderate | >15-20 | 9 | 14.2 | 2 | 3.3 |
| Moderate to severe | >20-25 | 5 | 7.9 | 1 | 1.6 |
| Severe | >25-40 | 0 | 0.0 | 0 | 0.0 |
| **Total** |  | **63** | **-** | **60** | **-** |

### World Health Organization-Five Well-Being Index (WHO-5)

The computed mean score at baseline (T1) was above the published threshold for positive wellbeing; the majority were above this threshold at both time points (39/47, 78.7% and 38/46, 82.6% respectively). Despite this mean score significantly increased post intervention (T2), (P=0.03) (Table 8).

**Table 8:** Table of comparison of mean wellbeing scores pre intervention (T1) and post intervention (T2); Mean scores, standard deviation (SD) and P-values calculated using a paired samples t-test.

| **Measurement** | **N** | **Mean<br>Pre** | **Mean<br>Post** | **SD<br>Pre** | **SD<br>Post** | **p-value** |
|---|---|---|---|---|---|---|
| WHO5 | 43 | 62.79 | 68.28 | 11.696 | 16.339 | 0.037 |

### Follow-up data (T3)

Only 7 participants (7/49, 14.2%) completed the three-month follow-up survey. At T3 mean OLBI score was 3.86 (SD, 5.273) range 0-15. All respondents met the criteria for ‘low levels’ of Burnout. At T3 mean CORE-10 score was 32.86 (SD, 7.010) range 22-41. The majority (6/7, 85%) were in the healthy range, and one met criterion for ‘moderate psychological distress’. At T3 mean WHO5 raw score was 20.57 (SD, 4.198) range 15-25. Again, above the published cut-off threshold for positive wellbeing. CORE-10 and WHO5 maintained statistically significant improvement at follow-up (Table 9).

**Table 9.** Comparison of mean scores for CORE-10, OLBI and WHO-5 at T1 and T3; Mean scores, standard deviation (SD), Z-scores and P-values calculated using Wilcoxen signed ranks.

| **Measurement** | **N** | **Mean T1** | **Mean T3** | **SD T1** | **SD T3** | **Z value** | **p-value** |
|---|---|---|---|---|---|---|---|
| OLBI | 7 | 39.06 | 32.86 | 5.031 | 7.010 | -1.527 | 0.127 |
| CORE-10 | 7 | 11.16 | 3.86 | 5.793 | 5.273 | -2.371 | 0.018 |
| WHO5 | 7 | 15.60 | 20.57 | 2.909 | 4.198 | -2.207 | 0.027 |

### Feasibility and acceptability and satisfaction

Programme fidelity was monitored throughout; All sessions were delivered as planned. The only changes implemented were for Cohorts 3 and 4, where the introductory session was delivered online. Daily attendance registers confirmed that all students who enrolled completed the full five-day programme.

The majority of participants were ‘very satisfied’ with the programme (41/62, 66.1%) and rated it ‘excellent’ (38/62, 61%), would recommend to others (58/60, 93.5%), and rated the instructors ‘excellent’ (47/62, 75.8%) (Table 10). Post intervention students were willing to incorporate holistic practices into their own lives and daily practice. Seventy one percent (46/49) stated that they believed the SKY holistic health intervention should be integrated into medical training and that they felt more equipped to use holistic approaches in patient care after taking part. Finally, all responded that they felt holistic practices could reduce drug dependency.

**Table 10:**
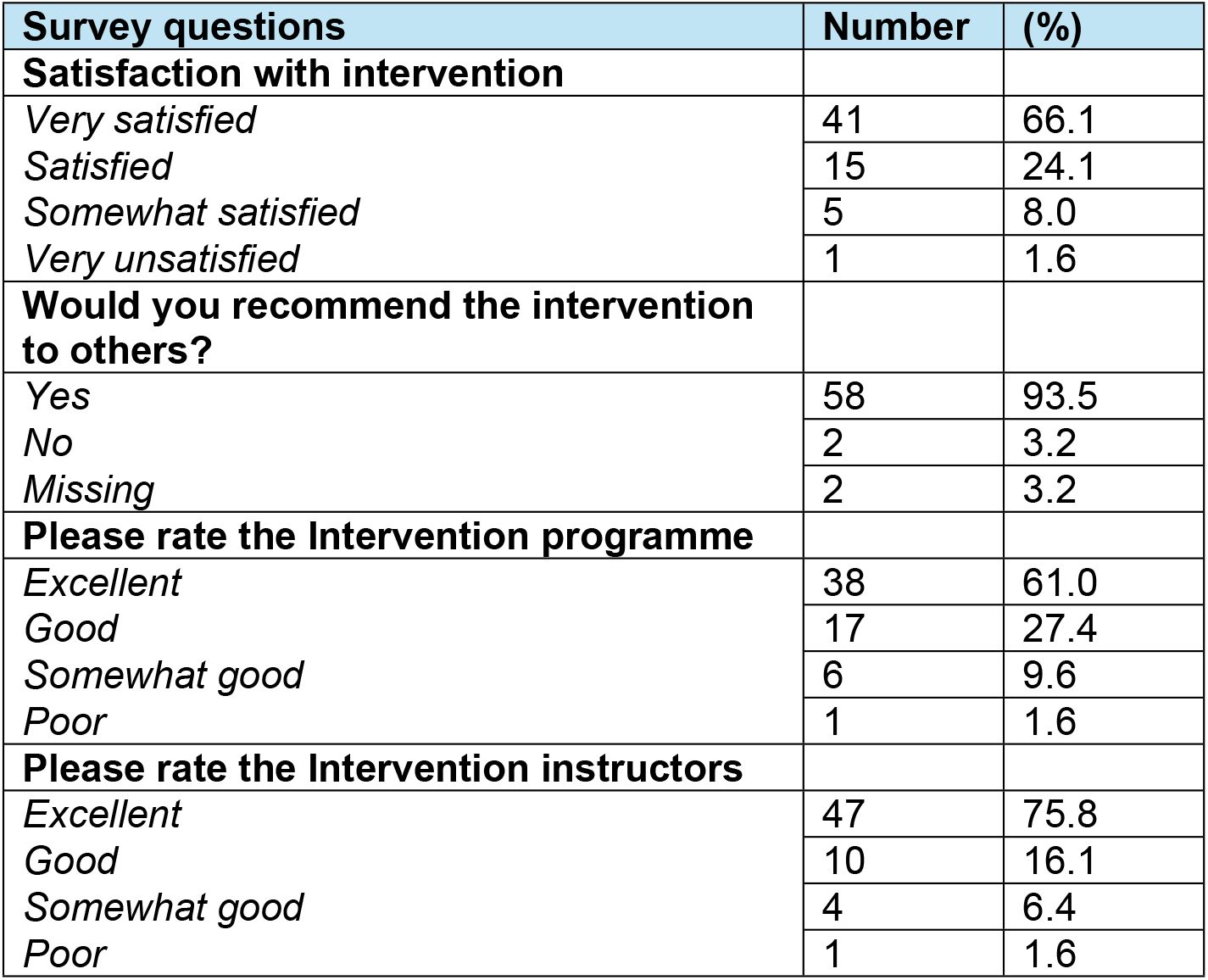
Frequency and percentage of satisfaction data across all four cohorts (n=62)

| **Survey questions** | **Number** | **(%)** |
|---|---|---|
| **Satisfaction with intervention** |  |  |
| *Very satisfied* | 41 | 66.1 |
| *Satisfied* | 15 | 24.1 |
| *Somewhat satisfied* | 5 | 8.0 |
| *Very unsatisfied* | 1 | 1.6 |
| **Would you recommend the intervention to others?** |  |  |
| *Yes* | 58 | 93.5 |
| *No* | 2 | 3.2 |
| *Missing* | 2 | 3.2 |
| **Please rate the Intervention programme** |  |  |
| *Excellent* | 38 | 61.0 |
| *Good* | 17 | 27.4 |
| *Somewhat good* | 6 | 9.6 |
| *Poor* | 1 | 1.6 |
| **Please rate the Intervention instructors** |  |  |
| *Excellent* | 47 | 75.8 |
| *Good* | 10 | 16.1 |
| *Somewhat good* | 4 | 6.4 |
| *Poor* | 1 | 1.6 |

### Free text comments

The T2 survey offered opportunity for free text comments; provided by 59.6% of participants (37/62). Comments reflected their enjoyment, perceived value, personal thank you messages to the instructions and suggestions for future delivery; namely shorter sessions and more experiential exercises. Participant quotes illustrate this (Table 11).

**Table 11:** Participant extract from T2 survey, free text comments.

| **Participant extract** | **Cohort** |
|---|---|
| *“I used breathing techniques to stay calm”*<br>*“I learnt lots of really interesting techniques, I am going to consider how I can implement them into my everyday life”* | Cohort 1 |
| *“The overall week was very helpful for me to discover many new techniques for relaxation. By trying new things every day it gave me flexibility to see what worked best for me which I liked”*<br>*“Thanks for such a great week, Dr Manmeet Kaur and Paul were very helpful, and I’ll be trying my best to keep following up with the techniques that I’ve learnt”* | Cohort 2 |
| *“Thank you very much! I truly believe Holistic Medicine should be embedded into all NHS policies”* | Cohort 3 |
| *“The instructors were really sweet and helpful throughout this week, thank you to all of them”*<br>*“Group sessions were really fun and interactive. All techniques were helpful and made a positive change on my life”* | Cohort 4 |

## Discussion

This study reports empirical data on the feasibility, acceptability and effectiveness of a five-day, in-person, MBI for burnout, psychological distress and wellbeing in UK undergraduate medical students. A statistically significant reduction in mean burnout (OLBI) post intervention (T2) was observed with a small effect size. A statistically significant reduction in mean CORE-10 scores post intervention was observed with a large effect size, maintained at follow-up. A statistically significant improvement in wellbeing (WHO5) was observed post intervention, with a small effect size, maintained at follow-up. Equally a statistically significant improvement post intervention was observed for current energy, stress, sustained focus and calmness.

Other literature exploring SKY breathing and mindfulness as a tool to reduce medical student burnout have shown varied results, with some finding significant reductions in burnout^23^, and others showing no change.^37,38^ This raises the question of how mindfulness interventions should be delivered to medical students to maximise benefit and tackle the multifaceted issue of burnout. These studies speculate that minimal change in burnout may have been due to short follow-up times, a lack of available students and limited engagement and adherence. Here the MBIs lasted eight and seven weeks respectively. This differs to the current intervention duration, which was much shorter; which may have been more impactful due to the time constraints and high workload commitments experienced by medical students.

The observed shift from ‘moderate’ to ‘mild’ psychological distress, post intervention suggests that the programme helped to improve medical students’ quality of life. These findings share similarities with current literature, demonstrating that MBIs can support stress management in medical students^39–41^. The practice of mindfulness encourages focus on the present moment^42^, with a non-judgemental awareness, making stress and anxiety feel more manageable.^43^ Supporting a reduction in psychological distress is essential to tackle the high prevalence of anxiety and depression within medical students^12^ and ensure a positive educational experience. Psychological distress has been associated with poorer academic self-perception^44^, higher dropout rates^45^, cynicism^46^ and decreased empathy.^47^ Therefore, brief interventions which reduce psychological distress are warranted to alleviate these issues.

Other research has also shown mindfulness to be effective in supporting subjective wellbeing in medical student populations.^48^ Importantly, medical students often face poorer wellbeing compared to their peers of the same age^12^; therefore, interventions which improve wellbeing are essential. The current population mean wellbeing scores were above the general population threshold for poor wellbeing but, despite this, an improvement was still observed. This is timely when studies report that medical students’ feelings towards current wellbeing interventions fall short, and act as more of a tick box exercise.^49^ In the report, limited engagement with interventions was attributed to time-tabling issues alongside clinical placement and inadequate advertisement. This may create a culture within medical education, in which students are discouraged from seeking mental wellbeing support. To combat this, mindfulness could be used to promote positive mental states within medical students. This intervention could act as an easily accessible wellbeing service that fosters greater discussion of mental health and wellbeing within the medical community, without impacting academia. Importantly, medical students who protect their wellbeing during their education may feel more able to provide compassionate and empathetic care, helping to improve doctor patient relationships in their future clinical practice.^50^

Acceptable interventions with high adherence are critical if medical curricular is to address these pressing issues. Free text feedback was highly positive, strengthened further by the 100% completion rate, this suggests the programme is acceptable to UK medical students when offered as a self-selected component of their medical training. This finding is further supported by the post intervention satisfaction data; the majority of participants across all four cohorts were highly satisfied with the programme, would recommend it to others and rated the programme and instructors as ‘excellent’. Equally the majority felt that the programme had made a difference to their life, found the techniques straight forward to do, and said that they would use the techniques in future when stressed. These findings were particularly important, as teaching people skills and techniques to cope with distress are an important aspect of mindfulness interventions. Thus, the current service evaluation adds empirical evidence to support the effectiveness and acceptability of MBIs in a UK medical student context.

This study also sought to explore medical students’ willingness to incorporate holistic practices into their own lives and into patient care. Participants reported a clear willingness to do so which implies a growing awareness among medical students around the benefits of mindfulness, both to improve wellbeing and patient care. In line with prior findings^51^, 88% of medical students felt there was a role for mindfulness as a tool for stress management, and 76% of students would consider referring patients for MBIs in the future. Interestingly, findings demonstrated more positive perceptions towards the application of mindfulness in mental healthcare, rather than physical healthcare. This suggests a need for awareness and education within medical students regarding the biological mechanisms by which mindfulness can provide benefits. The current MBI was successful in outlining the overall benefits of mindfulness, particularly through sharing evidence of biological processes. This may have contributed to the high satisfaction ratings and subsequent willingness to use holistic practices in participants. Therefore, when considering the implementation of mindfulness into medical curricula, we should perhaps consider educating medical students on the physiological mechanisms of this practice, as well as the wellbeing impacts of this practice.

This willingness to use mindfulness within daily life and patient care is also seen in healthcare staff who demonstrated an in-depth understanding of the benefits of mindfulness within a medical setting, such as through accepting present situations, fostering a shared humanity between colleagues and opportunity for self-care.^52^ Learning to accept present situations is essential with medical professionals, to help combat the emotional demands of their work with self-compassion.^53^ Similarly, increased communication within colleagues helps to improve the quality of patient care and foster a supportive work environment.^54^ A meta synthesis^52^ also highlighted some of the challenges of mindfulness training for healthcare staff, which included stigma around emotional vulnerability and associated persistent shame, and balancing work and life commitments. Such stigma could be described as an issue of work culture, whereby staff are discouraged to talk freely or seek support for mental health.^55^ Mindfulness is essential within this, self-awareness is critical requirement to foster the ability to recognise the need for self-care, alongside support between colleagues, could help to tackle this experience of shame. If mindfulness could be implemented at an early educational stage for medical students, this could help to alleviate stigma in professional training grades.

### Limitations

This study had some limitations. The sample was relatively small (n=69) and predominantly female, which may limit generalisability. However, research consistently suggests that female medical students face a higher risk of burnout^56^, driven by the unique interplay of several factors, such as heightened imposter syndrome^57^, perceived academic stress^58^ and vulnerability to emotional exhaustion.^59^ Nevertheless, it is important to ensure that when determining the effectiveness of interventions, that the sample is representative of the UK medical student population. In the UK, female students reportedly make up 60% of the population and in Northern Ireland even higher at 72% and so the current sample is similar. This female-majority trend among yearly intakes has been observed over several years and is now reflected in the wider workforce, where female doctors have reached parity with and overtaken men.^60^ So, it is imperative that evidence-based interventions that support females in particular, are evaluated and reported.

Wellbeing data was not collected in cohort 1 and so this sample size is smaller than the comparison made for the other outcome measures, again impacting generalisability. Equally, the response rate at 3 months was low (n=7) which limits the strength of any conclusions regarding longer-term impact. This is an important shortcoming to address in future studies.

Finally, the self-selected nature of the intervention participants may have created a risk of subjectivity^61^, in which those with an interest in Mindfulness or MBIs were biased towards a potential positive outcome. However, as mindfulness grows in popularity and becomes embedded in medical curricular this is likely to lessen the impact.

### Applications within Medical Education

A systematic review investigating mindfulness in medical schools found fourteen medical schools across the globe that teach mindfulness to medical students, dental students and residents.^62^ Similarly, nearly 80% of medical schools in the UK offer mindfulness activities, with five universities providing access to an eight-week wellbeing course, and seven universities embedding mindfulness as a required element in the core curriculum.^63^ Monash University have integrated mindfulness into their medical curricula with reduced psychological distress and improved quality of life during peak exam seasons^64^, implemented in multiple medical schools since.^63^ highlighting a clear interest and willingness to use mindfulness within medical education.

The format in which mindfulness is delivered currently differs between medical schools, including simple lectures, one-day workshops and two-month programmes. Considering this, it is also important to determine whether mindfulness should be taught as an elective or as part of the core medical curriculum. This idea has already been discussed^62^, raising the important point that there are obstacles and opportunities throughout the entirety of medical school. The most common way in which medical schools implement mindfulness is through a student selected process, implying that students are willing to engage. This motivation is arguably essential for student benefit, therefore, compulsory mindfulness in medical school may not benefit all.^62^ As a result, the present MBI should perhaps be introduced into Welsh medical schools as an optional component. Adjustments should be made to offer the current MBI at additional time points, to support students throughout their whole degree. Future qualitative research should focus on identifying student preferences for delivery of the SKY breathing HHC intervention in Welsh medical students, to maximise benefit. Similarly, future studies should also investigate the long-term impacts on Welsh medical students, to confirm whether these positive changes can be sustained.

## Conclusion

This study provides preliminary empirical data that supports the use of mindfulness-based interventions (MBIs) to promote positive wellbeing and improve treatment and management of burnout and psychological distress in UK undergraduate medical students. The five-day, in person Holistic Health Care (HHC) intervention incorporated SKY breathing techniques, Pranayama and yogic breathing, body scans and self-reflection. We report statistically significant improvements in burnout (OLBI), psychological distress (CORE-10) and wellbeing (WHO-5) with small to large effect sizes. The intervention programme had a significant and positive effect on participants self-reported energy and stress, sustained focus and calmness.

Completion rate was 100% and satisfaction data were highly positive. This suggests the programme is an effective and feasible approach to support to UK medical students’ mental health during their medical education journeys.

Future research should consider the implementation of the SKY breathing HHC intervention into Welsh medical curricula, as a student selected preventative tool to tackle burnout. It is also important to investigate student perspectives on how best to deliver the intervention and maximise impact. Long-term studies should be completed to establish the duration of effects, and whether taking part and receiving such training can support sustained improvements in mental health, particularly as students progress and become medical professionals working within a busy and complex national health service.

## Data Availability

The minimal data set is available UKdata share repository

## Notes

### Competing Interest Statement

I have read the journal's policy and the authors of this manuscript have the following competing interests: [Dr Manmeet Kaur is the Senior GP Partner, Pengam Health Centre, and CEO for Unity in Health Cymru CIC, Cardiff, Wales, United Kingdom. Manmeet was responsible for delivering the intervention to the students but was not involved in data analysis and data reporting.

